# Biases in routine influenza surveillance indicators used to monitor infection incidence and recommendations for improvement

**DOI:** 10.1101/2024.06.05.24308517

**Authors:** Oliver Eales, James M. McCaw, Freya M. Shearer

**Affiliations:** Infectious Disease Dynamics Unit, Centre for Epidemiology and Biostatistics, Melbourne School of Population and Global Health, The University of Melbourne, Australia; School of Mathematics and Statistics, The University of Melbourne, Australia; Infectious Disease Ecology and Modelling, Telethon Kids Institute, Perth, Australia

## Abstract

**Background:** Monitoring how the incidence of influenza infections changes over time is important for quantifying the transmission dynamics and clinical severity of influenza. Infection incidence is difficult to measure directly, and hence other quantities which are more amenable to surveillance are used to monitor trends in infection levels, with the implicit assumption that they correlate with infection incidence.

**Method:** Here we demonstrate, through mathematical reasoning, the relationship between the incidence of influenza infections and three commonly reported surveillance indicators: 1) the rate per unit time of influenza-like illness reported through sentinel healthcare sites, 2) the rate per unit time of laboratory-confirmed influenza infections, and 3) the proportion of laboratory tests positive for influenza (‘test-positive proportion’).

**Results:** Our analysis suggests that none of these ubiquitously reported surveillance indicators are a reliable tool for monitoring influenza incidence. In particular, we highlight how these surveillance indicators can be heavily biased by: the dynamics of circulating pathogens (other than influenza) with similar symptom profiles; changes in testing rates; and differences in infection rates, symptom rates, and healthcare-seeking behaviour between age-groups and through time. We make six practical recommendations to improve the monitoring of influenza infection incidence. The implementation of our recommendations would enable the construction of more interpretable surveillance indicator(s) for influenza from which underlying patterns of infection incidence could be readily monitored.

**Conclusion:** The implementation of all (or a subset) of our recommendations would greatly improve understanding of the transmission dynamics, infection burden, and clinical severity of influenza, improving our ability to respond effectively to seasonal epidemics and future pandemics.

## 1 Introduction

Influenza surveillance is important for managing the impact of seasonal influenza epidemics and future influenza pandemics. Measurements of how the incidence of influenza infections changes over time, such as a surveillance indicator that correlates with infection incidence, can be used to quantify the underlying transmission dynamics. In turn, measurements of the underlying infection levels can help quantify the clinical severity [1] of influenza infection, which may vary over time and by type, sub-type and age. With greater knowledge of the underlying transmission dynamics and clinical severity, forecasts and scenario projections can be improved, supporting improved public health interventions and ultimately improved population health outcomes.

The prevailing systems and methodologies for the surveillance of influenza epidemic activity have not changed substantially over more than three decades [2, 3, 4]. Routine influenza surveillance [5, 6, 7, 8] typically reports three surveillance indicators: the rate of influenza-like illness (ILI) reported through sentinel healthcare sites per unit time; the rate of laboratory-confirmed influenza (LCI) infections per unit time; and the proportion of laboratory tests positive for influenza (‘test-positive proportion’, TPP). These indicators, which are typically assumed to reflect trends in influenza infection levels, are often used to monitor trends in the infection incidence. However, there are often substantial differences between the trends of these indicators within and across influenza seasons (Figure 1) [9]. In such situations it is unclear which indicator(s), if any, reflect the underlying trends in the infection incidence.

**Figure 1:**
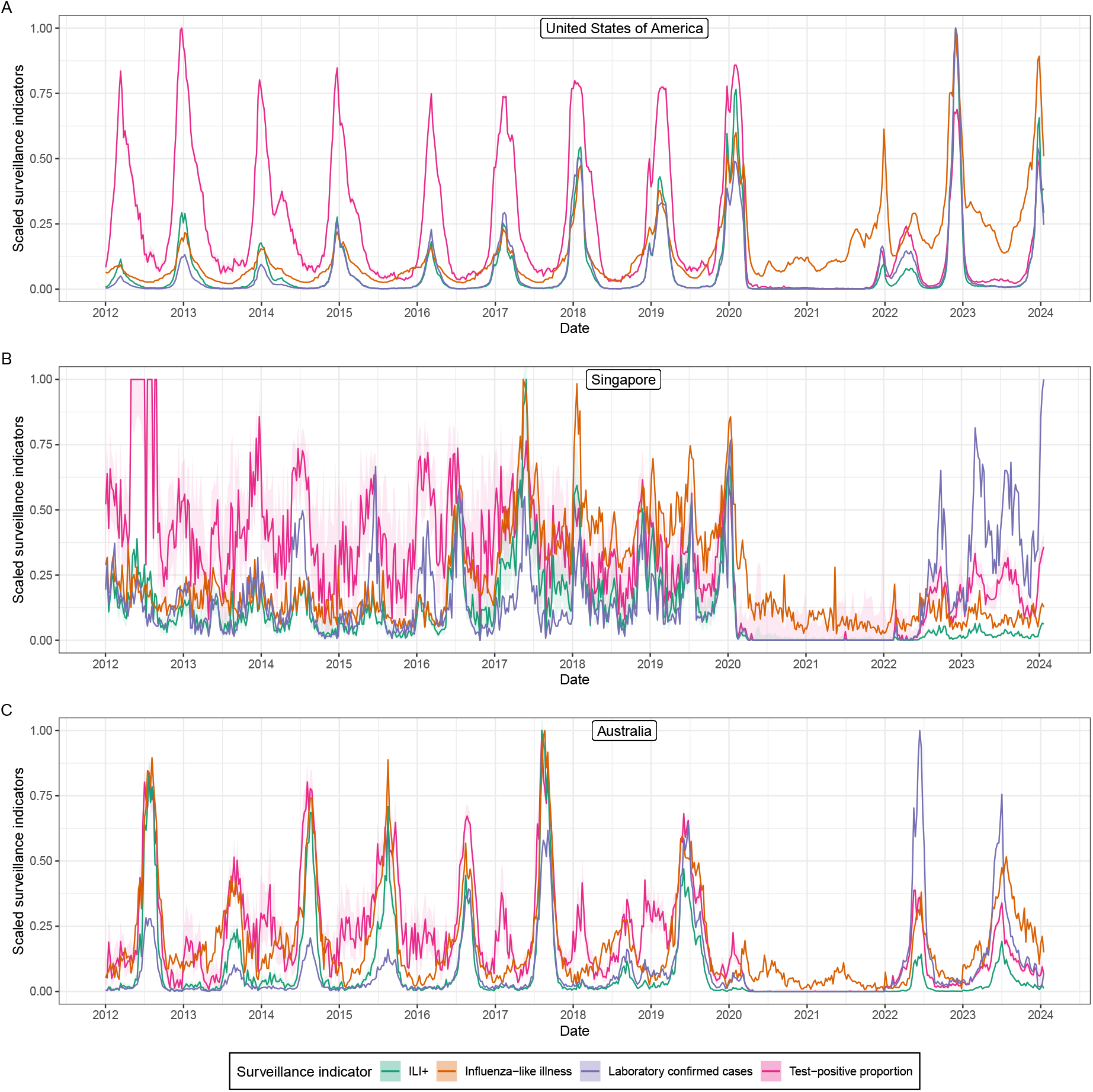
A comparison of surveillance indicators. Scaled influenza surveillance indicators from routine surveillance data collected in the United States of America, Singapore, and Australia [10] from the week starting 2 January 2012 to the week starting 15 January 2024. All surveillance indicators have been scaled so that their maximum value is one, to allow easier comparison of the time-series (the raw time-series are available in Figures S1, S2, and S3). The surveillance indicators shown are influenza-like illness (*ILI*(*t*), orange, see Section 2.1), laboratory confirmed influenza (*LCI*(*t*), purple, see Section 2.2), the test-positive proportion (*TPP* (*t*), pink, see Section 2.3), and *ILI*^+^(*t*) (green, see Section 3.2). 95% Binomial confidence intervals (shaded regions) are only shown for the *TPP* (*t*) and for *ILI*^+^(*t*); this is because the *TPP* (*t*) (which *ILI*^+^(*t*) depends on) was calculated from the total number of laboratory tests and the number of positive laboratory tests.

Here we demonstrate, through rigorous mathematical reasoning, how commonly reported influenza surveillance indicators relate to the incidence of influenza infections. We highlight how these relationships can be highly complex, time-dependent and arguably non-intuitive — for example, due to differences in behaviour and symptom rates by age, and due to the circulation of other respiratory pathogens. Given that the relationships between influenza incidence and these commonly reported surveillance indicators are so difficult to comprehend, it is critically important that we evaluate their interpretability and strive to develop more reliable indicators. We make six recommendations to improve public health surveillance for monitoring influenza infection incidence (Table 1). Aspects of our recommendations are already incorporated in WHO surveillance guidelines [4] and implemented in some countries [5, 6, 7, 8]. Furthermore, the WHO already recommends the routine collection of data required for most of our recommendations, and accordingly these data are collected by many countries. The implementation of our recommendations would require limited new investment in data collection and would enable the construction of surveillance indicators that are more readily interpretable due to their simple and well understood relationship to influenza infection incidence.

**Table 1:**
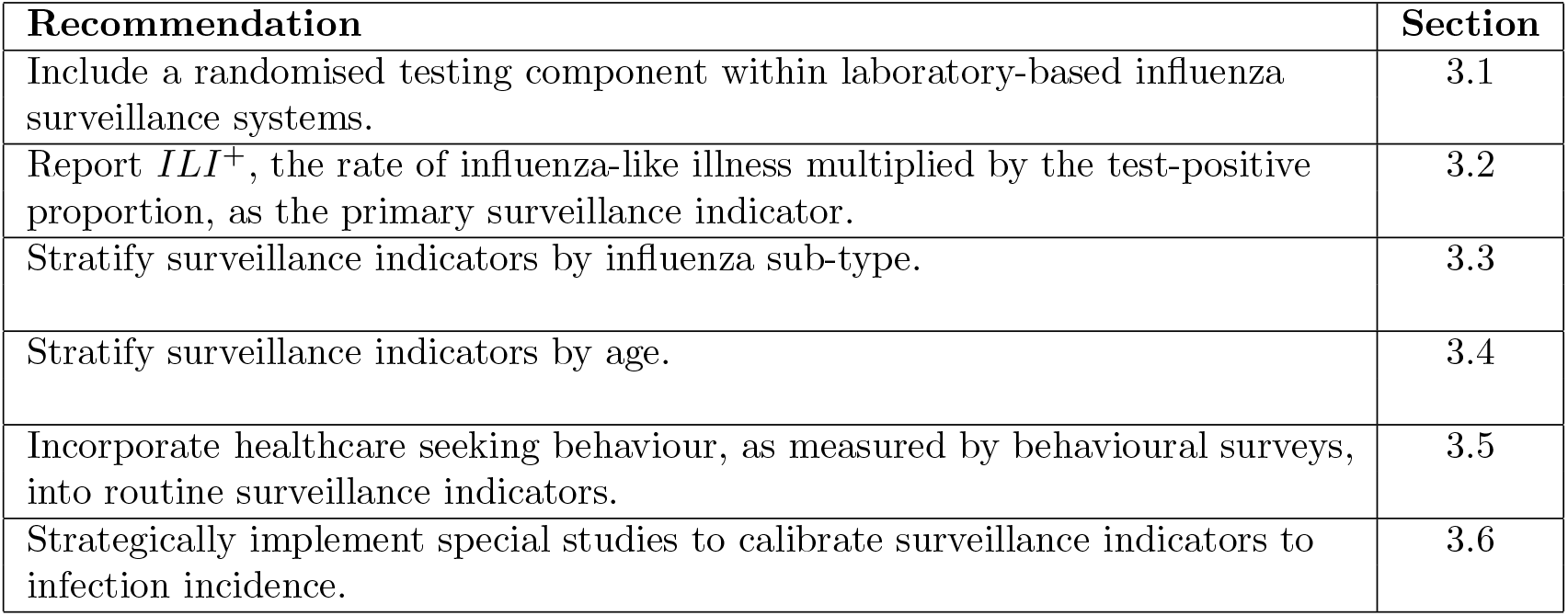
Recommendations to improve routine surveillance indicators and reporting for monitoring influenza infection incidence.

## 2 Biases in routine surveillance indicators

In this section, we describe the mathematical relationship between key influenza surveillance indicators and the incidence of influenza infections. Through these relationships we highlight the sources of bias that are present when using these routinely reported surveillance indicators for monitoring trends in influenza infection levels.

### 2.1 Influenza-like illness

Influenza infections represent only a small fraction of ILI and, because infection rates of influenza and other pathogens causing ILI vary through time asynchronously [11, 12, 13], the time-series of ILI is unlikely to reflect the temporal patterns in infection rates of any one of those pathogens (Figure 2). For example, during influenza seasons in the USA the proportion of ILI that is attributable to influenza infection (see test-positive proportion below) is highly variable and usually peaks at approximately 30% (Sup Fig. S1). In consequence, a time-series regularly used to quantify influenza dynamics mostly reflects the abundance of other pathogens (which also vary over time). This will not only bias trend analysis, but can also bias simple estimates like when influenza incidence begins to increase (i.e., the start of influenza season) which may be important for deciding on the timing of vaccination programs. For example in Figure 1, in the United States for multiple seasons from 2012-2020 the ILI curve increased from a baseline level earlier (by weeks to months) than ILI+ (see Section 3.2). In these situations, the early increase in the ILI indicator has obscured the beginning of the influenza season, with implications for vaccination program timing and other public health responses to the influenza season

**Figure 2:**
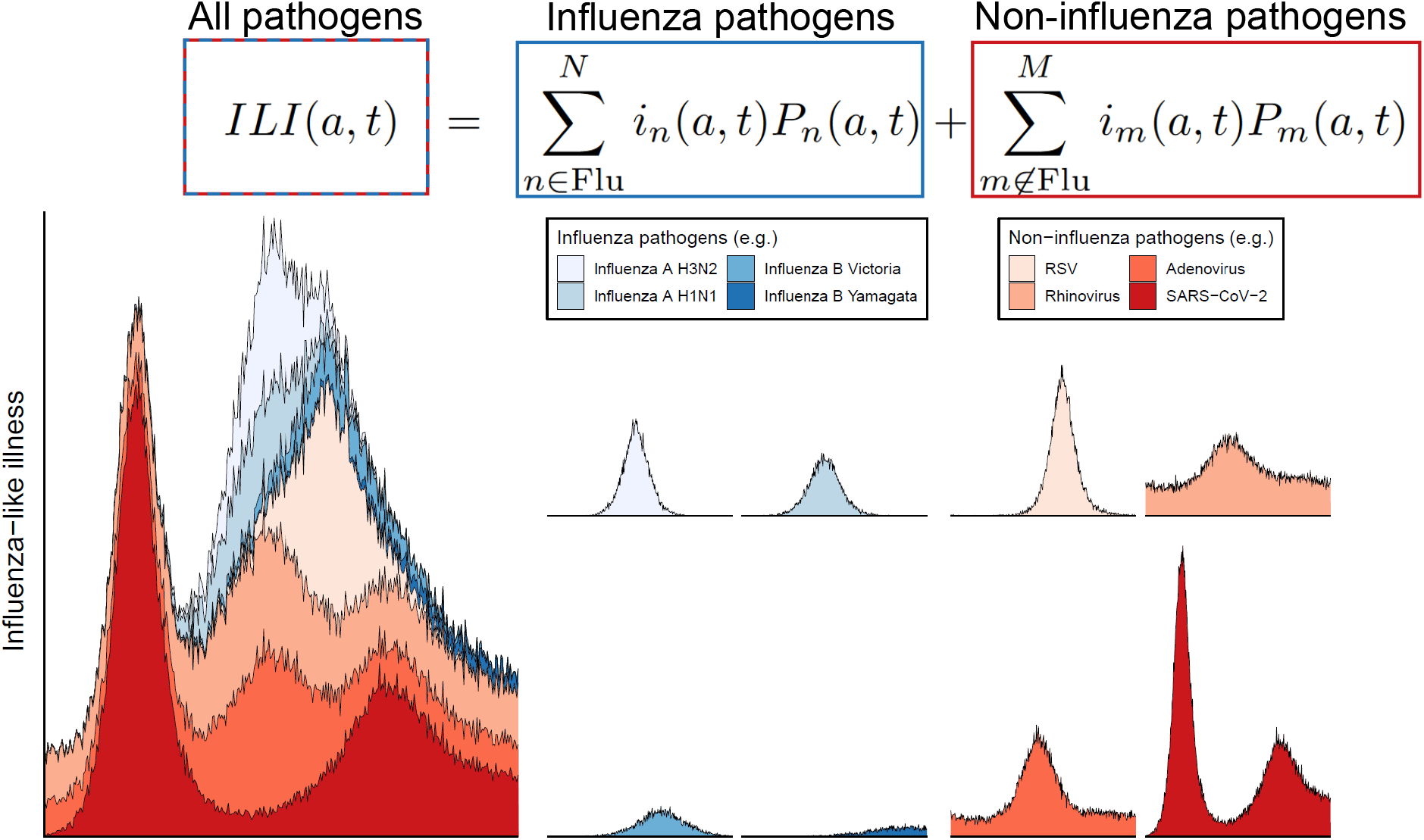
Graphical representation of the components of influenza-like illness. The time-series for the rate of influenza-like illness for individuals aged *a, ILI*(*a, t*), is made up of contributions from multiple pathogens that cause influenza-like illness. These pathogens can be influenza pathogens (blues) or non-influenza pathogens (reds). For each pathogen, its contribution to *ILI*(*a, t*) is dependent on the infection incidence (for individuals aged *a*) of the pathogen, *i*_*n*_(*a, t*) and the probability of an infected individual (aged *a*) being identified by a sentinel healthcare site. In general, there can be more pathogens contributing to *ILI*(*a, t*) than we have presented here. The time-series for all pathogen are only meant to be illustrative and so do not reflect actual trends as inferred from data.

If we assume that sentinel healthcare sites are consistent over time (e.g., same sites used, similar population coverage) the rate of ILI for individuals in a population aged *a* can be written as (see Figure 2 for visualisation):

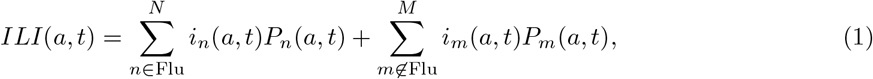

where *i*_*n*_(*a, t*) is the age-dependent infection incidence of a specific pathogen (*n*), and *P*_*n*_(*a, t*) is the age-dependent probability of an individual infected with pathogen *n* being identified by a sentinel site (i.e., the probability of an infected individual presenting to a healthcare facility and being recorded by the surveillance system). The summation applies to all pathogens that cause ILI-associated symptoms, but for clarity we separate it into two expressions: the left-hand expression sums over all influenza viruses (i.e., H3N2, H1N1, B) and the right hand expression sums over all other ILI-causing pathogens (e.g., SARS-CoV-2, respiratory syncytial virus). Note that we have simplified the expressions throughout by assuming that co-infection is sufficiently rare in the population that it can be neglected (for the purposes of estimating infection incidence).

In general, only the overall rate (as opposed to the age-dependent rate) of ILI in a population is reported. This is given by:

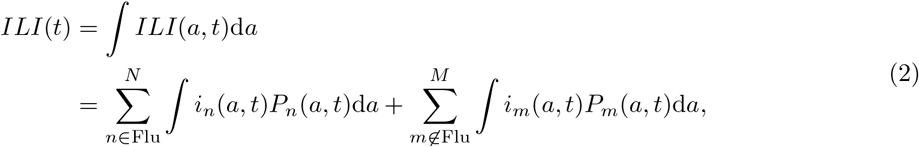

where we integrate the expression over all ages. This expression highlights that there will be clear biases in the time-series of *ILI*(*t*) where there are differences in epidemic dynamics and behaviour between age groups (see Section 3.4). The expressions for both *ILI*(*a, t*) and *ILI*(*t*) highlight that the time-series will be strongly dependent on other co-circulating pathogens with similar symptom profiles. Note that differences in transmission dynamics across spatial regions could be incorporated in a similar way to age. Sometimes ILI is reported as the percentage of visits to healthcare practitioners as opposed to the overall number [7]. This would introduce a time varying denominator to equation 2 describing the total number of visits to healthcare practitioners, rendering the surveillance indicator unnecessarily difficult to interpret.

### 2.2 Laboratory-confirmed influenza

Unlike ILI, Laboratory-confirmed influenza (LCI) will not depend on other circulating pathogens (assuming perfect test specificity), but it is influenced by the probability of laboratory testing being performed given presentation to healthcare with ILI, which can change over time (Figure S4). If we assume that tests for influenza have perfect specificity (no false positives), then the age-dependent rate of LCI can be written as:

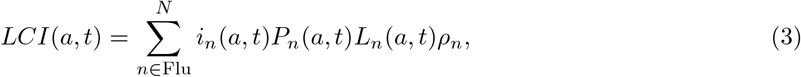

and the overall rate of LCI can be written as:

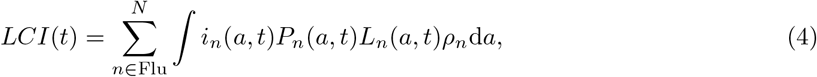

where *L*_*n*_(*a, t*) is the probability that a laboratory test is ordered for an individual who has sought healthcare with ILI due to infection with pathogen *n*, and *ρ*_*n*_ is the sensitivity of the test for pathogen *n*. The probability of a laboratory test being ordered by a health practitioner can vary over time (both between seasons and within a single season) introducing time-dependent effects, and may differ by age (introducing further age effects) and between pathogens due to differences in the perceived severity of an infection (see Section 3.1).

### 2.3 Test-positive proportion

The test-positive proportion (often referred to as the “percentage positive”) is, despite its prominence in surveillance reports, arguably the most peculiar, prone to bias, and difficult to interpret surveillance indicator used to represent influenza infection incidence. The age-dependent proportion of laboratory tests positive for influenza can be written as:

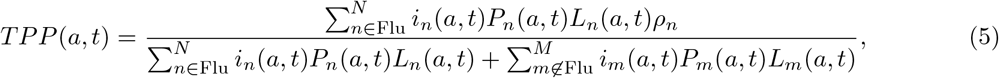

and the overall proportion of laboratory tests positive for influenza is given by:

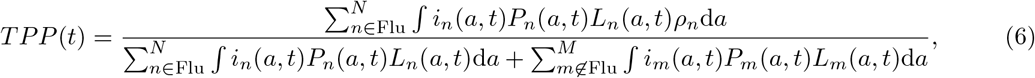

where the numerators are the same as equations 3 and 4, and the denominator is simply the total number of laboratory tests performed (either by age group or overall). If we assume that no other ILI-causing pathogens are circulating (i.e.,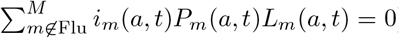) then the value of equation 6 is only dependent on the sensitivity of the tests for identifying the different sub-types of influenza (*TPP* (*t*) = *ρ*, when sensitivity is the same across sub-types). The test-positive proportion measures the infection incidence of influenza relative to the infection incidence of other pathogens (which are also unknown). Increases in *TPP* (*t*) can reflect increases in influenza incidence or decreases in the incidence of other pathogens (e.g., SARS-CoV-2). Comparing peaks in the test-positive proportion between influenza seasons to comment on differences in influenza infection levels is therefore fraught — for example, compare the 2012–2013 and 2019–2020 flu seasons in the USA (Figure 1). The peak in the test-positive proportion in the 2012–2013 influenza season was the highest, but all other surveillance indicators reached far greater values in the 2019–2020 influenza season (greater than twice the values reached in the 2012–2013 flu season). Similarly, when increases (and decreases) in influenza incidence occur synchronously (and at a similar magnitude) to increases in the incidence of other ILI-causing pathogens, we expect *TPP* (*t*) to be approximately constant despite the transient increases in influenza incidence.

## 3 Recommendations to improve routine surveillance indicators

All three surveillance indicators that we have considered are dependent on multiple unknown and unmeasured quantities, rendering their interpretation risky. By developing surveillance indicators that do not depend on these unknown or unmeasured quantities, we will gain new insight into the underlying infection incidence of influenza. To that end, we make six practical recommendations for improving influenza surveillance. Some aspects of the recommendations are already recommended by the World Health Organization (WHO) [4], but even when they are implemented they are not properly integrated into routine reporting. Rather, the (unrepresentative) surveillance indicators described above are still far more widely reported. We show sequentially how each of our recommendations removes a layer of complexity from the (mathematical) description of the relationship between surveillance indicator and infection incidence. The result is a simplified and interpretable surveillance indicator(s) from which underlying patterns of infection incidence could easily (and safely) be inferred.

### 3.1 Include a randomised testing component within laboratory-based influenza surveillance systems

The probability that an influenza test is ordered for an individual (presenting with ILI at a healthcare facility) can depend on decisions made by the healthcare provider, which may vary over time. Infections perceived to be more severe might have a greater probability of undergoing testing. This can introduce differences in the probability of testing depending on the causative pathogen — for example, SARS-CoV-2 infections can present as influenza-like illness, but may also include other symptoms [14] perceived to be more severe. Additionally, there can be age-specific differences in the probability of testing — the elderly and young children are at the greatest risk of severe influenza infections [15] and so may be more likely to be tested. If testing to confirm influenza infection was performed randomly on patients presenting with ILI (*L*_*n*_(*a, t*) = *L*(*t*) for all *n*) the *LCI*(*t*) surveillance indicator is a more accurate reflection of infection incidence. This is demonstrated by the simplification of equation 4 to:

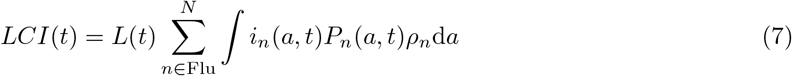

where *L*(*t*) is no longer inside the integral. We note that random testing of ILI presentations would need to be performed in parallel to testing for clinical purposes. For example, testing every Nth individual presenting with influenza-like illness, ‘interval sampling’, is a straightforward method that is recommended by the WHO [4]. In performing any downstream analysis of infection incidence, it would be important to only use the randomly selected samples; a small random sample is more informative than a larger biased sample.

If the probability of testing, *L*(*t*), was constant over time, then the expression for *LCI*(*t*) (equation 7) could be further simplified. However, this is not necessary (see below) and may not be preferential as sample sizes may not be sufficient during certain epidemic periods if the sampling probability was constant over time.

### 3.2 Report *ILI*^+^ as the primary surveillance indicator

‘*ILI*^+^’ — the rate of influenza-like illness, *ILI*(*t*), multiplied by the test-positive proportion, *TPP* (*t*) [16] — is a more meaningful surveillance indicator, and will more closely reflect trends in the underlying infection incidence (relative to the routinely reported surveillance indicators described above). When a random component of laboratory testing to confirm influenza infection is performed (*L*_*n*_(*a, t*) = *L*(*t*) for all *n*), i.e., in parallel to clinical testing, the expression for *ILI*^+^ is given by:

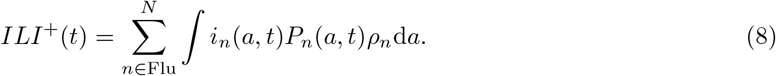

This is similar to the expression for the rate of lab-confirmed influenza, but without the dependence on the rate of laboratory testing (which can change over time). Although the relationship between *ILI*^+^(*t*) and infection incidence is still complex (depending on differences in infection dynamics and the probability of an individual being identified by ILI surveillance over time, by age, and by influenza sub-type), it is more interpretable (and less prone to bias) compared to the other surveillance indicators considered as it has no dependence on other circulating pathogens (like *ILI*(*t*) and *TPP* (*t*)), nor does it depend on laboratory testing rates (like *LCI*(*t*)); there can be substantial differences between the time-series of *ILI*^+^(*t*) and these other surveillance indicators (Figure 1). There are examples of *ILI*^+^(*t*) being used in the academic literature to quantify influenza infection dynamics [16, 17, 18], but *ILI*^+^(*t*) is rarely reported as a part of routine surveillance [5, 6, 7, 8], despite being a simple composite of routinely reported surveillance indicators that we have rigorously demonstrated is a more reliable and interpretable indicator of influenza infection incidence. Additionally, if laboratory testing can also identify when the etiological agent of ILI is an ILI-symptom causing pathogen other than influenza (e.g., SARS-CoV-2, parainfluenza, rhinovirus), then analogous surveillance indicators can be constructed for each pathogen i.e., existing influenza surveillance strategies [19] could be used to infer transmission dynamics of many more pathogens.

### 3.3 Stratify surveillance indicators by influenza sub-type

Influenza sub-types exhibit different epidemic dynamics. For example, the timing and frequency of epidemics of influenza A and B can vary substantially (Figure 3). The age-profile of epidemics can also be different with a greater proportion of younger age infections for influenza B relative to influenza A and in influenza A H1N1 relative to influenza A H3N2 [20]. Additionally, there may be sub-type specific differences in the probability of infected individuals presenting to healthcare facilities (e.g., due to differences in symptom rates). The time-series of *ILI*^+^ does not distinguish between different influenza sub-types and so cannot distinguish these different epidemic dynamics. But making this distinction is important for forecasting future epidemic activity, especially as different influenza sub-types can interact at an epidemiological scale [21], and also for forecasting the rate of clinical outcomes as influenza sub-types can differ in their severity.

**Figure 3:**
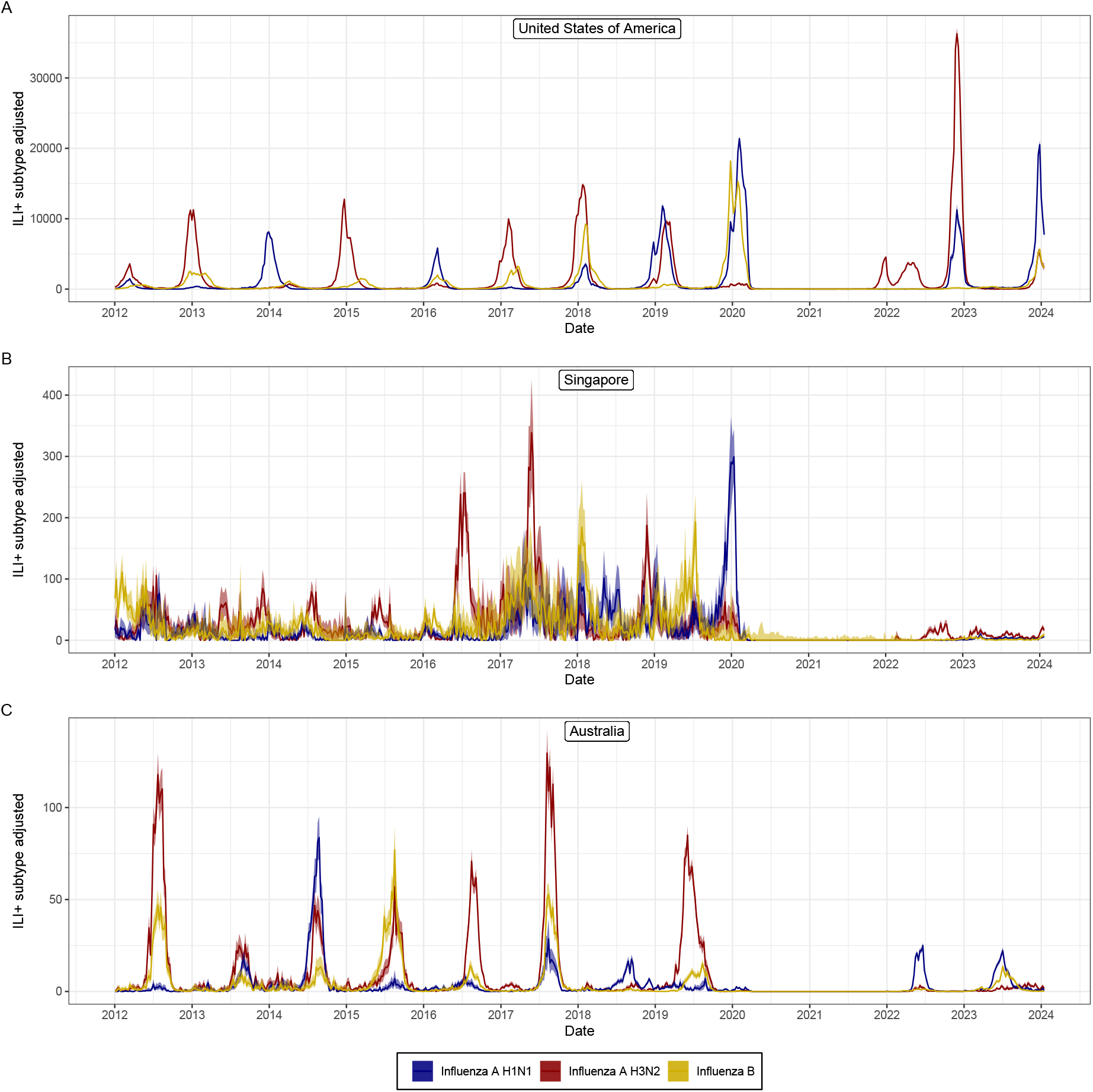
Sub-type specific values of. *ILI*^+^(*t*) Sub-type specific values of *ILI*^+^(*t*) as estimated from routine surveillance data collected in the United States of America, Singapore, and Australia [10] from the week starting 2 January 2012 to the week starting 15 January 2024. For all time-series we have plotted the central point estimate (line) and the 95% confidence interval (shaded region). We have considered only influenza A H3N2, influenza A H1N1, and influenza B (any lineage). Each time-series was calculated by multiplying the rate of influenza-like illness, *ILI*(*t*), by the test-positive proportion (the number of positive laboratory tests divided by total number of tests), *TPP* (*t*), and by the proportion of each specific sub-type. The proportion of influenza B was simply the number of specimens determined to be influenza B divided by the total number of specimens for which influenza type could be determined (B, A H3N2, A H1N1, A unsubtyped). To estimate the proportion of A H3N2 and A H1N1 we first estimated the proportion of influenza A (H3N2, H1N1 and unsubtyped), the same as above for influenza B, and then multiplied it by the proportion of each influenza A sub-type (calculated from the specimens in which a sub-type could be determined).

In many countries a subset of laboratory-confirmed influenza cases are also tested to determine the influenza sub-type [5, 6] as recommended by the WHO [4]. If these data are collected (by random sampling of laboratory confirmed influenza cases) the relative proportion of each influenza sub-type contributing to *ILI*^+^(*t*) (or other surveillance indicators) can be estimated (Figure 3). Then the surveillance indicators will only be dependent on the influenza sub-type of interest, with each indicator more accurately reflecting the respective individual sub-type dynamics. We can then obtain the sub-type specific value of *ILI*^+^, 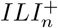 (for influenza sub-type *n*), by multiplying *ILI*^+^(*t*) with the relative proportion of sub-type

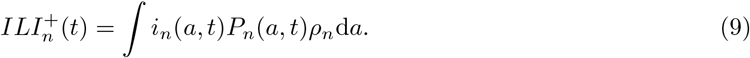

There are examples in the academic literature of data on sub-type proportions being used to stratify surveillance indicators, including *ILI*^+^(*t*), in order to better quantify the epidemic dynamics of a single influenza sub-type [22, 23]. However, this is rarely done for routine surveillance outputs, where instead only the number of positive specimens for each sub-type [5] or the test-positive proportion for each sub-type [6] might be reported, neither of which has a clear epidemiological interpretation. Additionally, routine surveillance outputs will typically report the daily number (or proportion) of influenza A H3N2, influenza A H1N1, and influenza A unsubtyped (specific sub-type is not always determined) specimens (and influenza B, etc). This further obscures any differences in the dynamics between H3N2 and H1N1 and could be accounted for by using the relative proportion of H3N2 and H1N1 (confirmed) specimens to estimate their relative proportion in the influenza A unsubtyped specimens.

### 3.4 Stratify surveillance indicators by age

We have until now ignored the complexity introduced to surveillance indicators due to differences between age-groups. Surveillance indicators used to describe epidemic activity are often reported for the overall population (i.e., not age-stratified). Differences between age groups are known sources of bias in surveillance indicators [4], and there is often an implicit (incorrect) assumption in public health surveillance that this bias is constant through time. However, the surveillance indicator 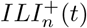 depends on both the age- and time-dependent incidence, *i*_*n*_(*a, t*), and age- and time-dependent probability of presenting at healthcare (given infection), *P*_*n*_(*a, t*) (equation 10). If there is no interaction between the age- and time-dependence for both quantities (e.g., if the infection incidence can be split into a time-varying component and age-varying component: 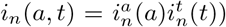 then the bias due to differences between age-groups will be constant through time. This is demonstrated by the simplification of equation 10 to:

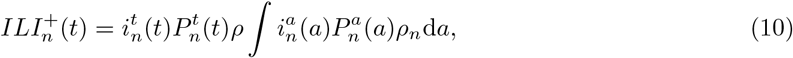

where the integral term on the right is now simply a constant that scales the expression. If there is no interaction between age- and time-dependence then these (non age-stratified) surveillance indicators would be justified — the time-series may over represent specific age-groups, but trends over time would be the same. However, interactions between time- and age-dependence are common for both quantities: relative levels of infection incidence between age groups can change through time (see [24] for example with SARS-CoV-2 prevalence, and Figure S5 for example using age-stratified ILI data); and differences between age groups in the probability of presenting for healthcare can change over time [23]. Therefore, biases due to differences between age-groups are expected to vary through time.

Age-effects could be accounted for if age were recorded with all influenza surveillance data (as recommended by the WHO [4]) and surveillance indicators were then stratified by age. In some countries, age is collected with influenza surveillance records, allowing for age-stratification [5]. Age-stratified surveillance indicators will more accurately reflect trends in (age-stratified) influenza infection incidence. The age-stratified values of 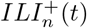 (equation 10) can be written as:

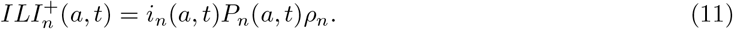

Each age-stratified surveillance time series is directly proportional to the infection incidence and the probability of presenting at healthcare (given infection) for the specific age group. In reality, there will still be differences in infection dynamics across the age range of a single age group; the greater the number of age groups used to stratify the indicator, the more accurate the representation will be of the underlying transmission dynamics. While small sample sizes may render visualisations of the raw data for highly stratified indicators less informative, appropriate statistical methods can be applied to quantify the expected (mean) trends in stratified indicators and their associated uncertainty.

### 3.5 Incorporate behaviour surveys into routine surveillance indicators

The probability than an infected individual is identified and recorded by a surveillance system is impacted by behavioural factors, including whether an individual seeks healthcare, which in turn may be influenced by socio-demographic and physical environmental factors. To correctly infer how the (age-stratified) infection incidence (of a specific influenza sub-type) varies over time from routine influenza surveillance data, the probability of an infected individual presenting for healthcare (*P*_*n*_(*a, t*)) — and how it varies over time — must be quantified (see equation 11). The probability of an individual presenting for healthcare (when infected with a given influenza sub-type) can be decomposed into two components: the probability an infected individual develops influenza-like symptoms, *S*_*n*_(*a, t*); and the probability an individual with influenza-like symptoms seeks healthcare, *H*_*n*_(*a, t*). With this decomposition, equation 11 can be written as:

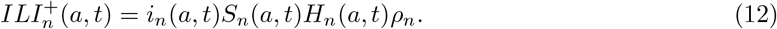

Without estimates of how these two functions change over time, estimates of temporal trends in infection incidence will be biased. Changes over the time-scale of a single season, will bias the estimated epidemic dynamics, affecting the inferred shape of the epidemic curve, including peak timing, size, and sharpness. Changes over longer time-scales will bias estimates of the relative infection levels between seasons. Symptom surveys that also collect data on whether individuals sought healthcare can be used to quantify healthcare seeking behaviour (i.e., *H*_*n*_(*a, t*)) [25, 26]. Healthcare seeking behaviour can be explicitly incorporated into surveillance indicators, generating ‘healthcare seeking behaviour adjusted’ surveillance indicators, [23] as follows:

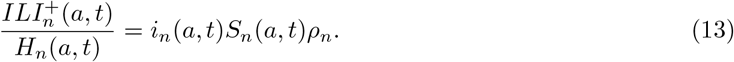

Participatory surveillance systems that collect these data already exist in many countries (e.g., “Flu-Tracking” in Australia [27], “Outbreaks Near Me” in the USA [28]). Although the individuals who participate in such surveys may reflect a more health conscious subset of the population [25], it is likely that trends over time and by age are broadly consistent (i.e., the estimated probability of seeking health-care may be skewed, but by a constant degree).

### 3.6 Strategically implement special studies to calibrate surveillance indicators

Special studies, such as infection prevalence studies (where a representative sample of the population is tested for influenza infections) [29, 30, 31] could be used to accurately estimate infection incidence (without the biases associated with routine influenza surveillance). When symptom surveys are included in infection prevalence studies, the probability that an infected individual develops influenza-like symptoms (*S*_*n*_(*a, t*)) — a key quantity for understanding the underlying infection burden from routine influenza surveillance data — can also be directly estimated. The probability that an infected individual develops symptoms may change over the time-scale of a single season (e.g., background symptom rates may increase in winter months), and it is highly likely that there are changes between seasons (e.g., new strains may be more or less virulent) and so estimates would need to be made periodically. Infection prevalence studies are not currently used for routine influenza surveillance because they are costly. However, they could be used in a small number of jurisdictions or a specific cohort (e.g., employees of a large business and their households) at strategic time points to estimate the probability of developing symptoms as a function of age and time (i.e, *S*_*n*_(*a, t*)); there is likely large agreement in this quantity between countries and jurisdictions, as symptom rates are a biological factor (as opposed to behavioural) and population level prior exposure rates are likely similar (as immunological profiles may also affect symptom rates). The jurisdictions that invest in such studies would benefit the most, with the greatest understanding of local transmission dynamics and the ability to calibrate their time series of surveillance indicators against direct estimates of infection incidence (at snapshots in time). While there are benefits to making calibration factors and quantities (such as the probability of symptoms) estimated for a jurisdiction widely available, the value of their application to another context will depend on the similarity in epidemiology and surveillance systems between the two jurisdictions.

## 4 Conclusion

We have described (using a precise mathematical formulation) the relationships between influenza infection incidence and three routinely reported influenza surveillance indicators (TPP, ILI, LCI). Our analysis suggests that none of these indicators are a reliable tool for monitoring the epidemic activity of influenza. In particular, we have demonstrated that the relationship between TPP and infection incidence is especially opaque and prone to bias. Furthermore, ubiquitously reported ILI (and TPP), is strongly influenced by the dynamics of circulating pathogens with similar symptom profiles — with their interpretability (and hence utility) further compromised in recent years by the emergence of SARS-CoV-2 [32].

We provide six recommendations for how routine influenza surveillance indicators used to monitor infection incidence could be improved. We recommend reporting *ILI*^+^ as the primary surveillance indicator, using data from a randomised testing component within laboratory-based influenza surveillance systems. We recommend stratifying all surveillance indicators by influenza sub-type and age. Finally, we recommend supplementing routine surveillance with behavioural surveys (e.g., through participatory surveillance systems) and periodic special studies (e.g., infection prevalence studies) to calibrate surveillance indicators to infection incidence. We expect that the implementation of these recommendations will be highly feasible since the collection of required data are already recommended by the WHO [4], and accordingly, many countries routinely collect these data [5, 6, 7]. However, on surveillance *reporting*, WHO advises countries to report a number of surveillance indicators which we have demonstrated are prone to bias [4]. Furthermore, *ILI*^+^, which we have demonstrated is a better reflection of infection incidence, is not included in WHO reporting guidelines. Hence, a review of surveillance reporting guidelines is warranted.

The implementation of all (or a subset) of our recommendations would greatly improve understanding of the transmission dynamics, infection burden, and clinical severity of influenza, improving our ability to respond effectively to seasonal epidemics and future pandemics.

## Data Availability

All data used in the present work are publicly available and cited in the manuscript

## Supplementary figures

**Figure S1:**
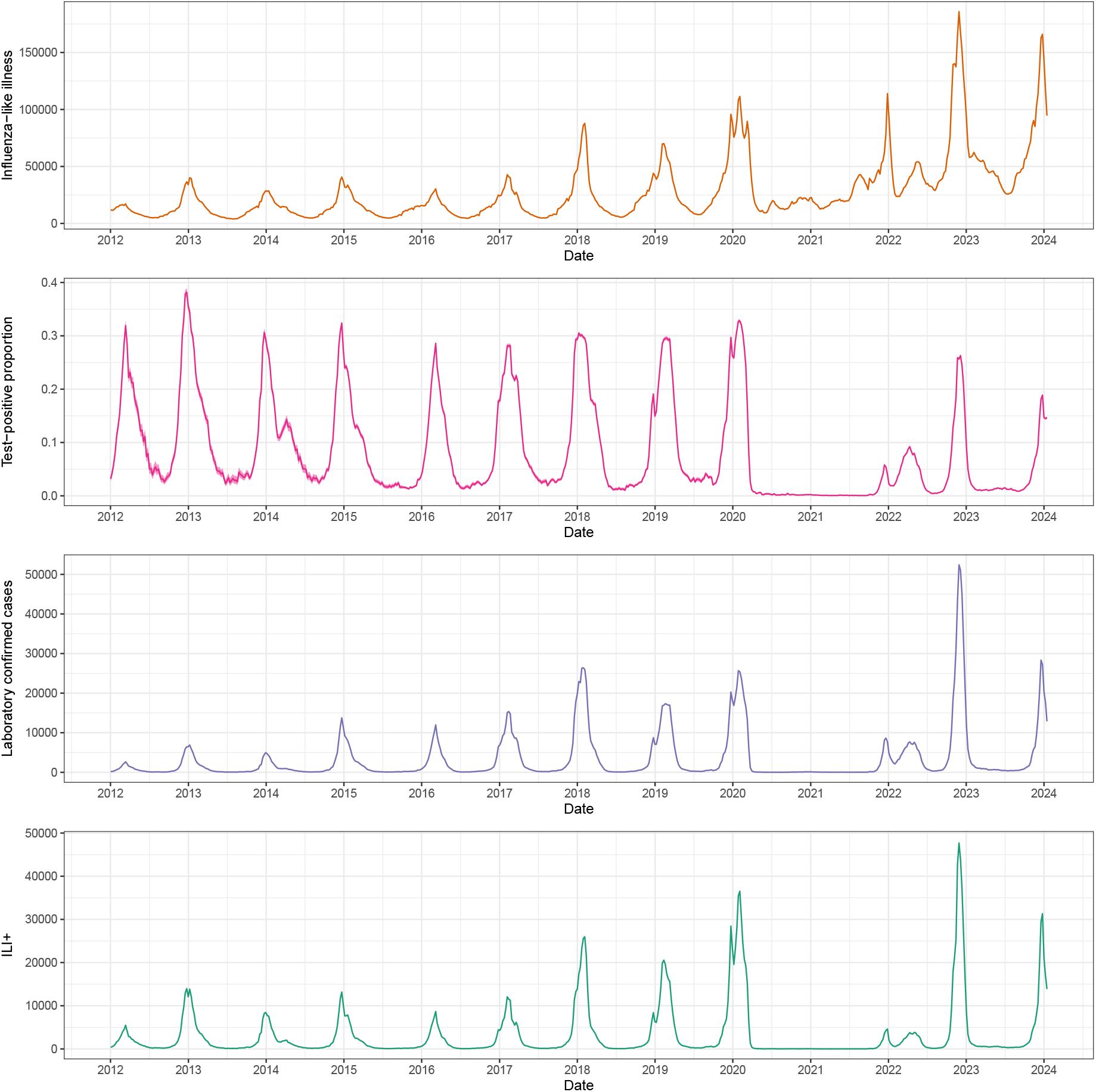
Time-series of surveillance indicators in the United States of America. The time-series of surveillance indicators from routine surveillance data collected in the United States of America [10] from the week starting 2 January 2012 to the week starting 15 January 2024. The surveillance indicators shown are influenza-like illness (*ILI*(*t*), orange, see Section 2.1), laboratory confirmed influenza (*LCI*(*t*), purple, see Section 2.2), the test-positive proportion (*TPP* (*t*), pink, see Section 2.3), and *ILI*^+^(*t*) (green, see Section 3.2). 95% Binomial confidence intervals (shaded regions) are shown for the test-positive proportion and for *ILI*^+^(*t*); this is because the test-positive proportion (which *ILI*^+^(*t*) depends on) was calculated from the total number of laboratory tests and the number of positive laboratory tests.

**Figure S2:**
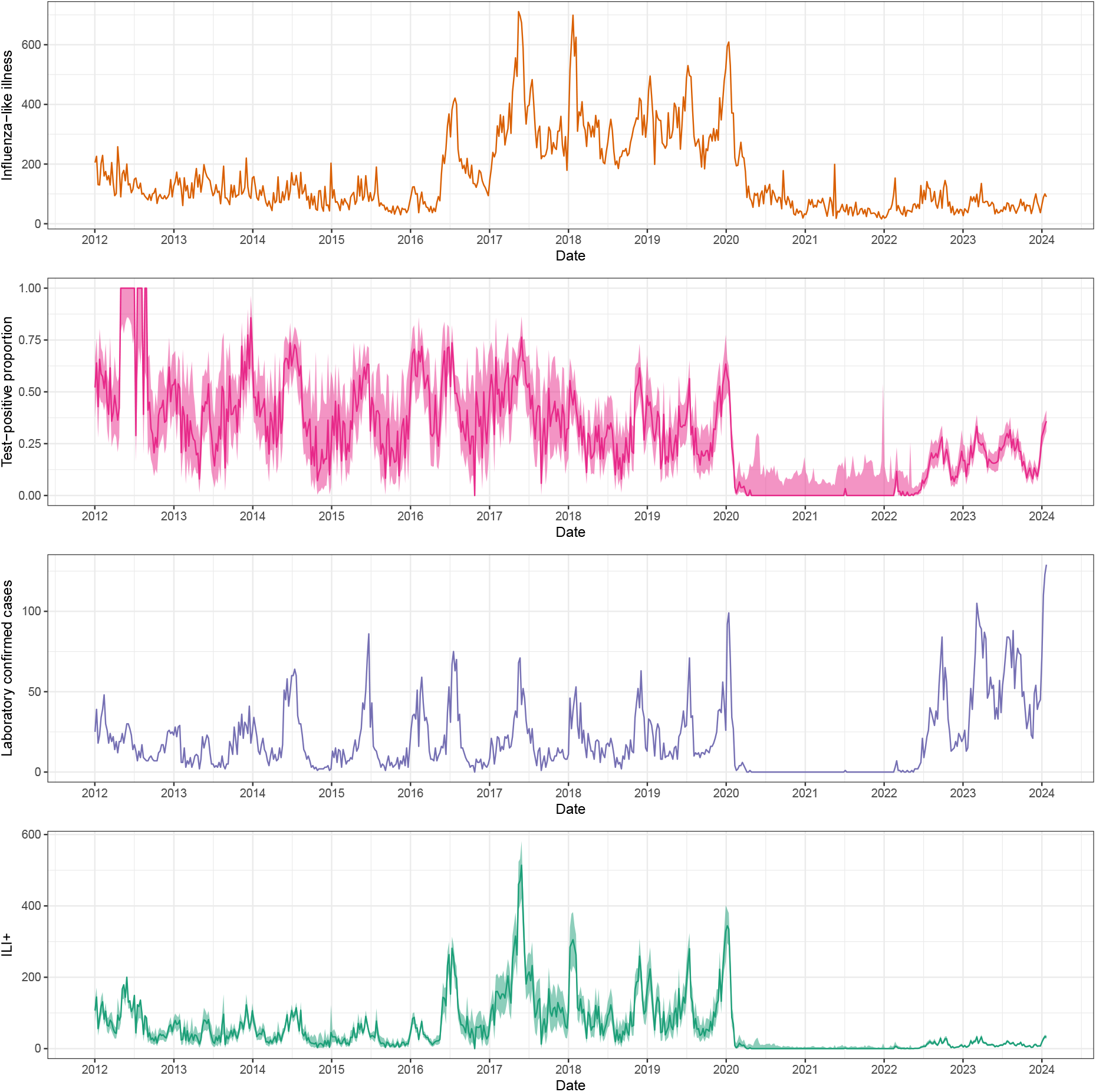
Time-series of surveillance indicators in Singapore. The time-series of surveillance indicators from routine surveillance data collected in Singapore [10] from the week starting 2 January 2012 to the week starting 15 January 2024. The surveillance indicators shown are influenza-like illness (*ILI*(*t*), orange, see Section 2.1), laboratory confirmed influenza (*LCI*(*t*), purple, see Section 2.2), the test-positive proportion (*TPP* (*t*), pink, see Section 2.3), and *ILI*^+^(*t*) (green, see Section 3.2). 95% Binomial confidence intervals (shaded regions) are shown for the test-positive proportion and for *ILI*^+^(*t*); this is because the test-positive proportion (which *ILI*^+^(*t*) depends on) was calculated from the total number of laboratory tests and the number of positive laboratory tests.

**Figure S3:**
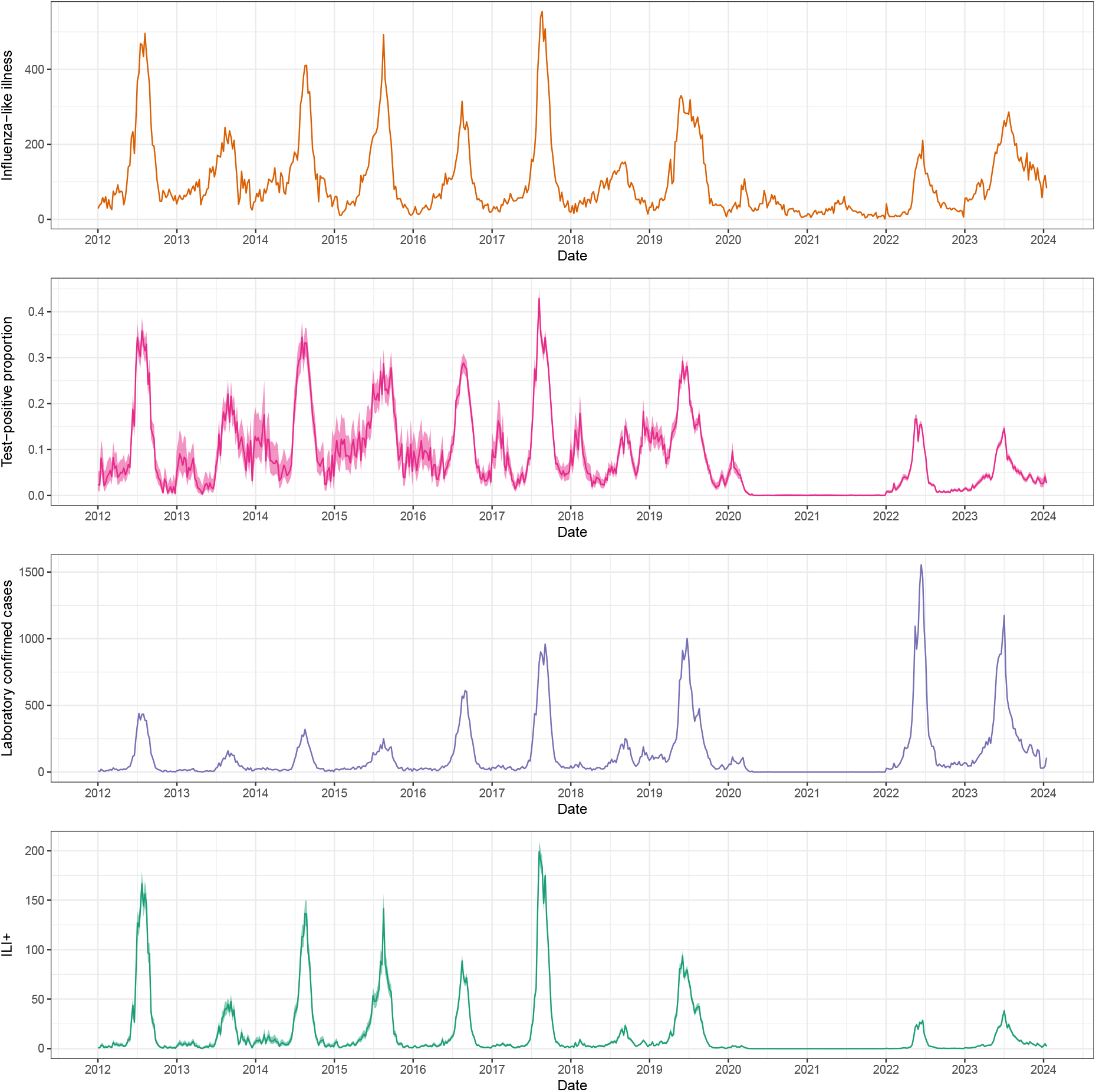
Time-series of surveillance indicators in Australia. The time-series of surveillance indicators from routine surveillance data collected in Australia [10] from the week starting 2 January 2012 to the week starting 15 January 2024. The surveillance indicators shown are influenza-like illness (*ILI*(*t*), orange, see Section 2.1), laboratory confirmed influenza (*LCI*(*t*), purple, see Section 2.2), the test-positive proportion (*TPP* (*t*), pink, see Section 2.3), and *ILI*^+^(*t*) (green, see Section 3.2). 95% Binomial confidence intervals (shaded regions) are shown for the test-positive proportion and for *ILI*^+^(*t*); this is because the test-positive proportion (which *ILI*^+^(*t*) depends on) was calculated from the total number of laboratory tests and the number of positive laboratory tests.

**Figure S4:**
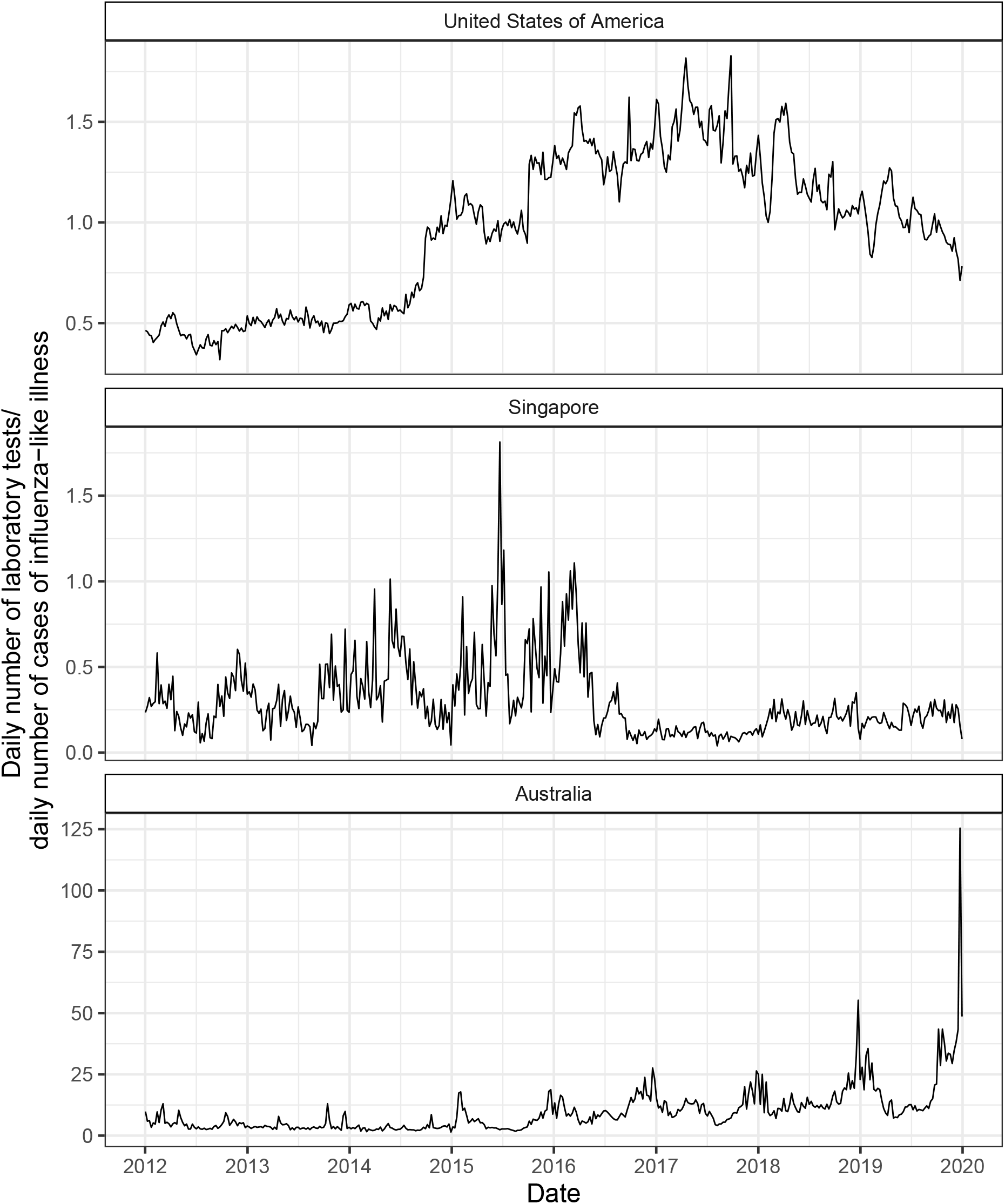
Ratio of laboratory testing to influenza-like illness. The ratio of the daily number of laboratory tests to the daily number of cases of influenza-like illness for data collected in the United States of America, Singapore, and Australia [10] from January 2012 to December 2019. This ratio should correlate with the probability that a laboratory test is ordered for an individual exhibiting influenza-like illness who has sought healthcare. Note that cases of influenza-like illness are identified through sentinel surveillance sites, whereas the number of laboratory tests is not limited to sentinel surveillance sites, and so the number of laboratory tests can be greater than the number of cases. We have only included data up to 2020 to demonstrate typical variation in the probability of a laboratory test being ordered (should correlate with the ratio plotted) across typical influenza seasons.

**Figure S5:**
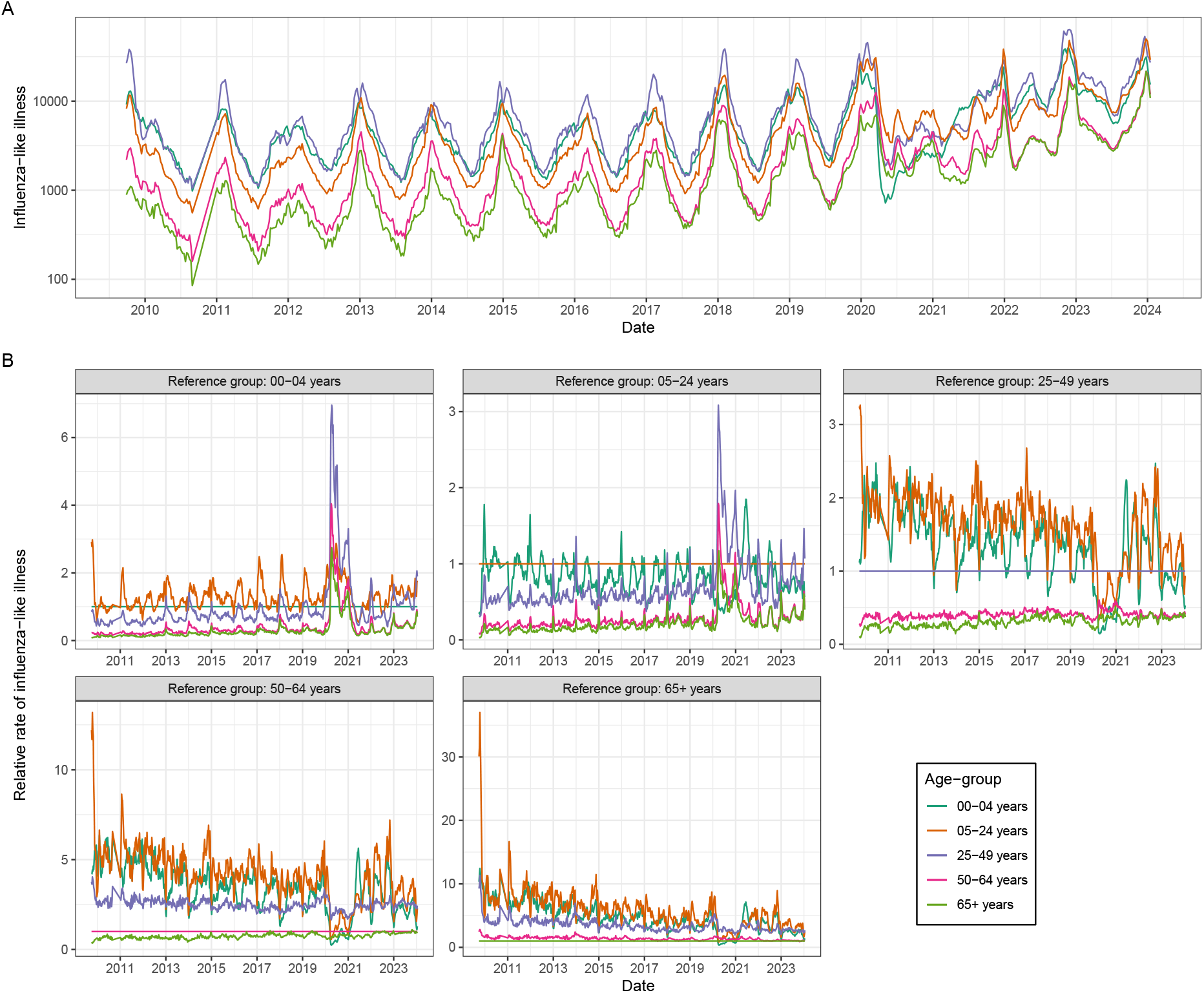
Differences in the temporal trends of influenza-like illness between age-groups. (A) The time-series of influenza-like illness across five age-groups (0-4 years, 5-24 years, 25-49 years, 50-64 years, 65+ years) from routine surveillance data collected in the United States of America [10] from the week starting 28 September 2009 to the week starting 15 January 2024. Note that the y-axis is on a log10 scale to allow easier comparison of the time-series. (B) The time-series of influenza-like illness for each of the five age-groups divided by the time-series of influenza-like illness for of the other age-groups (reference group). The reference group is different for each panel. This visualisation highlights that the relative difference in the rate influenza-like illness between age-groups changes over time.

